# Empirical Frequency Bound Derivation Reveals Prominent Mid-Frontal Alpha Associated with Neurosensory Dysfunction in Fragile X Syndrome

**DOI:** 10.1101/2022.02.24.22271485

**Authors:** Ernest V Pedapati, John A. Sweeney, Lauren M. Schmitt, Lauren E. Ethridge, Makoto Miyakoshi, Rui Liu, Elizabeth Smith, Rebecca C. Shaffer, Steve W. Wu, Donald L. Gilbert, Paul S. Horn, Aaron Buckley, Craig A. Erickson

## Abstract

The FMR1 gene is inactive in Fragile X syndrome (FXS), resulting in low levels of FMRP and consequent neurochemical, synaptic and local circuit neurophysiological alterations in the fmr1 KO mouse. In FXS patients, electrophysiological studies of have demonstrated a marked reduction in global alpha activity and regional increases in gamma oscillations that have been associated with intellectual disability and sensory hypersensitivity. Since alpha activity is associated with thalamocortical function that has widely distributed modulatory effects on neocortical excitability, insight into alpha physiology may provide insight into systems-level disease mechanisms. Herein, we took a data driven approach to clarify the temporal and spatial properties of alpha and theta activity in participants with FXS. High-resolution resting-state EEG data was collected from participants affected by FXS (n=65) and matched controls (n=70). We used a multivariate technique to empirically classify neural oscillatory bands based on their coherent spatiotemporal patterns. Participants with FXS demonstrated: 1) a redistribution of lower-frequency boundaries indicating a “slower” dominant alpha rhythm, 2) an anteriorization of alpha frequency activity, and 3) a correlation of increased individualized alpha power measurements with auditory neurosensory dysfunction. These findings suggest an important role for alterations in thalamocortical physiology for the well-established neocortical hyper-excitability in FXS, and thus a role for neural systems level disruption to cortical hyperexcitability that has been studied primarily at the local circuit level in mouse model research.

## Introduction

The FMR1 gene is inactive in Fragile X syndrome (FXS) which results in reduced Fragile X Mental Retardation Protein (FMRP) production and varying levels of intellectual disability, autistic characteristics, and sensory hypersensitivity (Baker et al., 2019; Hagerman et al., 2017). Despite a considerable understanding of molecular and microcircuit alterations in the *Fmr1*^-/-^ KO(Dahlhaus, 2018), systems-level changes in human brain and their relationship neurosensory function and behavior remain poorly understood in FXS. EEG studies have become increasingly used in the FXS field, both in patients and mouse models, to provide insight into central disorder mechanisms at the level of brain systems (Ewen et al., 2019; Zhang et al., 2018). We and others have identified reproducible, group-level abnormalities in human resting-state EEG and auditory evoked potentials in FXS which are associated with intellectual disability, neuropsychiatric symptoms, and sensory hypersensitivity(Ethridge et al., 2019; Ethridge et al., 2016; J. Wang et al., 2017). These changes include 1) a global reduction in alpha power (8-12 Hz) and increase in theta power (3.5-7.5 Hz) and 2) an increase in background gamma power (>30 Hz) (Ethridge et al., 2019; Ethridge et al., 2016; Musumeci et al., 1988; Van der Molen & Van der Molen, 2013; J. Wang et al., 2017; Wilkinson & Nelson, 2021).

In humans, alpha activity including a dominant peak frequency of approximately 10 to 13 Hz (upper alpha) has specific functional relevance across several major neurophysiological systems in typically developing individuals (Bollimunta et al., 2011; Fries, 2015; Garcia-Rill et al., 2016; Jensen & Mazaheri, 2010; Zhang et al., 2018). As a radio receiver prefers specific electromagnetic frequencies, neurons and oscillatory networks also demonstrate frequency preferences (Tseng et al., 2014; Tseng et al., 2006). In several neuropsychiatric disorders, including epilepsy, schizophrenia, neuropathic pain, and tinnitus, a lowering of the dominant peak frequency from the alpha to theta range has been observed (Vanneste et al., 2018). Furthermore, in these conditions, a lower frequency power in the theta range has been more strongly associated with elevations in gamma power. This constellation of findings, a lowering of the dominant frequency toward the theta range with parallel increases in resting gamma activity has been referred to as thalamocortical dysrhythmia (TCD, Lakatos et al., 2020; Llinas et al., 2005). In animal models of TCD, hyperpolarization of thalamic relay neurons has been observed to lead to alterations in firing patterns of low-threshold calcium spikes which are reflected in local field potentials as elevated theta rhythmicity(Jeanmonod et al., 1993; Jeanmonod et al., 2003; Llinás et al., 1999). A dominant theta rhythm may be reflective of a diminished inhibitory capacity at a systems-level resulting in excess asynchronous gamma activity and the so-called gamma “edge effect” (Golovin & Broadie, 2017; Llinás et al., 1999; Paluszkiewicz et al., 2011; R et al., 2005).

In FXS, we have hypothesized that a dominant theta peak frequency may be insufficient to drive neural ensembles with an alpha preference in thalamocortical circuitry and the canonical role of alpha in establishing inhibition-timing windows for optimal sensory and neurocognitive processing(Bonnefond & Jensen, 2015; Jensen et al., 2014; Jensen & Mazaheri, 2010; Zhang et al., 2018). At present, it remains an open question in FXS whether alterations in low-frequency activity represent two distinct pathologies (e.g., elevated theta and reduced alpha activity) or a single process (i.e., reducing alpha frequencies into what is typically considered the theta frequency band). To study this question, we used a data-driven approach to classify resting-state neural activity to compare FXS and matched control participants. We employed a “bottom-up” approach of classifying empirical bands of EEG activity distinguished by their coherent spatial and temporal features(Cohen, 2021). We reasoned that if alpha activity was “slowed” in FXS the total number of boundaries of empirical bands would remain the same as controls, but the spatiotemporal patterns within these bands would be altered. Conversely, if the EEG patterns in FXS represent either a consolidation of bands (i.e., a single theta band) or additional bands (i.e., novel spatiotemporally distinct alpha bands), then there would be a difference in the number of detected empirical bands than controls.

We tested these hypotheses in a large cohort of individuals with FXS and sex and age matched controls using high-resolution resting-state EEG. Following an empirical reconstruction of spatiotemporal boundaries (**see Figure 1**) that we found that changes in the alpha band in FXS were more consistent with a pervasive shift (mediated by genotype), rather than a reduction or increase of patterns of brain activity. The findings shed light into the nature of alpha and theta oscillations in FXS including clinical relevance and implicates a larger role for thalamocortical dysfunction.

**Figure 1:**
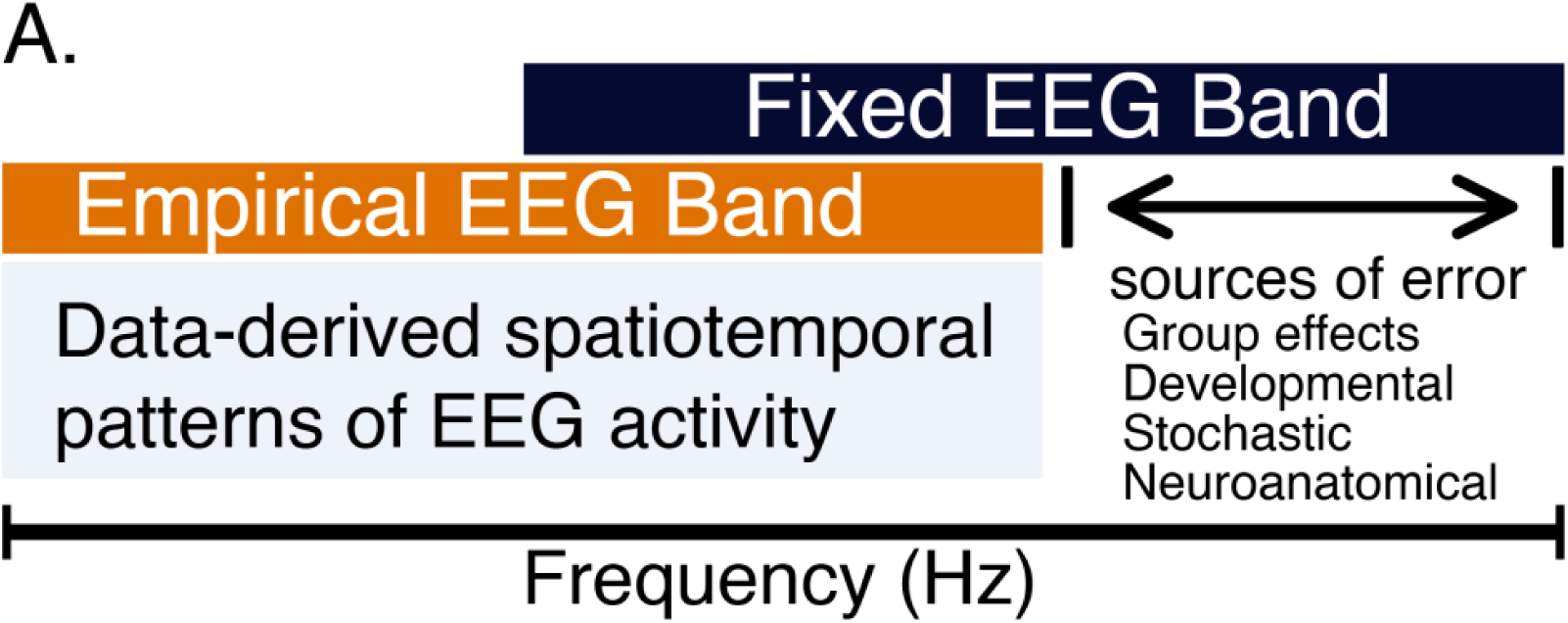
Schematic. A. Utility of data-driven classification of spatiotemporal frequency boundaries in neurodevelopmental disorders in precision medicine applications.

## Results

### Participants

The final dataset of resting-state EEG data contained 65 participants with FXS (37 males) and 70 healthy controls (41 males) with demographic and clinical characteristics in **Table 1**. Cases and controls did not differ in age or sex ratio. Participants with FXS had reduced full scale intelligence quotient (IQ), verbal IQ (VIQ), non-verbal IQ (NVIQ) and increased impairment in social communication and symptoms of anxiety. NVIQ is a general measure of intellectual capacity which is useful when verbal ability varies among individuals as in FXS. EEG preprocessing was blinded to group assignment, and no group differences in epoch number, number of interpolated channels, or ICA-derived artifact components were found (**see Supplemental Table 1**).

**Table 1:** Participant characteristics by group and sex with mean (± standard deviation) group significance testing. FXS, Fragile X Syndrome; y, years; FSIQ, Full Scale IQ; NVIQ, Non-verbal intelligence quotient; VIQ, verbal intelligence scale; Social Score, Social Communication Questionnaire; Anxiety Score, Anxiety, Depression and Mood Scale Anxiety Inventory (ADAMS).

|  | **FXS(F)** | **FXS(M)** | **Control(F)** | **Control(M)** | **p** |
| --- | --- | --- | --- | --- | --- |
|  | **N=28** | **N=37** | **N=29** | **N=41** |  |
| Age (y) | 19.3 (9.48) | 22.2 (9.61) | 22.2 (12.2) | 22.2 (9.81) | 0.609 |
| FSIQ | 64.7 (28.8) | 34.5 (23.2) | 99.9 (6.94) | 106 (10.1) | <0.001 |
| VIQ | 71.4 (26.9) | 46.8 (24.6) | 97.3 (9.42) | 108 (13.3) | <0.001 |
| NVIQ | 58.0 (33.2) | 22.1 (28.9) | 103 (8.65) | 104 (12.2) | <0.001 |
| Social Score | 9.12 (6.52) | 17.4 (6.39) | 2.75 (2.57) | 1.73 (1.93) | <0.001 |
| Anxiety Score | 5.65 (4.68) | 7.69 (5.00) | 2.41 (3.16) | 2.03 (2.22) | <0.001 |

### Exemplar of empirical bound detection in a single subject

For each subject, a series of channel covariance matrices were generated from the artifact-free EEG data to succinctly model spatiotemporal relationships for a generalized eigenvalue decomposition (GED). The detection of a series of empirical bands for a single subject is shown in **Figure 2**. The heat plot depicts the strength of spatiotemporal correlation (r^2^) between a series of narrow-band frequency channel covariance matrices (S_MATRIX_) and broadband channel covariance matrix (R_MATRIX_). The block-diagonal appearance of the shaded boxes represents the degree of r^2^ of spatiotemporal coherent EEG activity; adjacent boxes demonstrate the distinct spatiotemporal dynamics of distinct activity bands. The magenta line surrounding each shaded box are statistically significant clusters detected by a *dbscan* cluster algorithm. At the bottom of the heatmap, a blue line plot depicts the normalized magnitude of separation (the top eigenvalue per frequency step) between the S_MATRIX_ and R_MATRIX_. Adjacent to each cluster is a topographical plot of principal component analysis (PCA) averaged eigenvectors from each S_MATRIX_ and R_MATRIX_ pair which represents a dipole-like projection of the empirical band of spatiotemporal coherent neural activity. Subject-level derivation plots for all 135 subjects are available in the supplemental materials.

**Figure 2:**
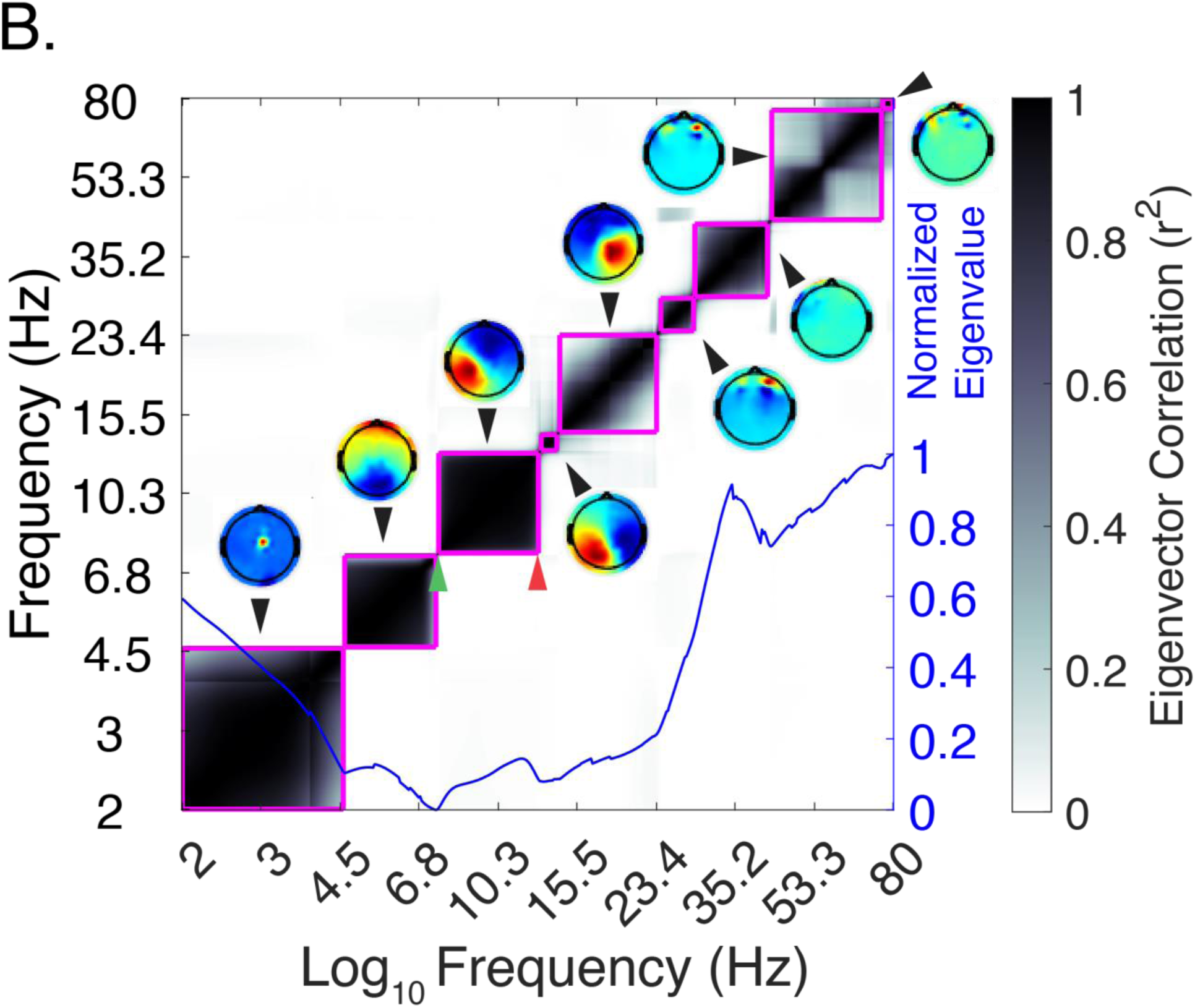
Single subject empirical bounds exemplar. Heat plot depicts strength of spatiotemporal linear correlation coefficient between narrow frequency filtered and broadband signal channel co-variance matrices (see Methods). Coherent blocks of distinct patterns of neural activity are delineated by a clustering algorithm (magenta line). Each block contains a lower (green arrow) and upper (red arrow) detected boundary. Topographic source projects constructed from PCA-averaged eigenvectors are place adjacent to heatmap clusters. The magnitude of the separation between narrow and broadband matrices is represented by eigenvalues (blue line).

### Group-level summary of empirical bound detection results

#### Number of detected bands

We first tested if the number of identified empirical EEG bands varied between the groups. The number of detected empirical bands of coherent spatiotemporal activity between 2 and 80 Hz did not differ (mean ± standard deviation) between FXS (M: 12.5±3.4; F: 12.0±3.6) and control (M:12.5±3.8; F: 10.8±3.1; **see Table 2**). No difference in number of detected bands below 30 Hz between FXS (M: 9.27±2.9; F: 8.8±2.5) and controls (M: 9.5±3; F: 8.5±2.8) was present.

**Table 2:** Summary of detected frequency boundaries. Group comparison of data-driven frequency boundaries mean (± standard deviation).

|  | **FXS(F)** | **FXS(M)** | **Control(F)** | **Control(M)** | **p** |
| --- | --- | --- | --- | --- | --- |
|  | **N=28** | **N=37** | **N=29** | **N=41** |  |
| Below 30 Hz | 8.79 (2.47) | 9.27 (2.91) | 8.48 (2.82) | 9.51 (3.00) | 0.442 |
| Above 30 Hz | 3.25 (1.84) | 3.19 (1.66) | 2.28 (1.33) | 2.95 (1.67) | 0.091 |
| 2-80 Hz | 12.0 (3.61) | 12.5 (3.43) | 10.8 (3.08) | 12.5 (3.76) | 0.177 |

#### Density of detected band boundaries

Next, we estimated the continuous density of the upper and lower frequency boundaries of the detected empirical bands within each group using a kernel density estimator (KDE). A higher KDE value indicated that many participants in the group shared a boundary at a specific frequency. We predicted that the density of frequency boundaries was shifted in FXS and that this difference varies based on sex and by frequency. We ran a linear mixed effect model (LME) in R (version 4.1) using the *nlme* package. We included KDE as the dependent variable and added fixed effects of group, sex, boundary frequency, as well as interaction effects. Subject was included as a random effect. A three-way interaction effect was significant (Lower: F_77,10087_=1.72, p=.005; Upper: F_77,10087_=1.3, p=.047; **see Figure 3**). Post-hoc comparisons identified significant sex-matched group differences primarily within the canonical theta and alpha frequency ranges (**see Supplemental Table 2**). Both males and females with FXS displayed a greater density of boundaries within the theta band (3.5-6 Hz) but differences, as expected, were more pronounced in the male subgroup. In FXS, the peak density for both lower and upper bounds occurred in the lower theta band then remains relatively uniform until an increase in density in the high alpha band (11-12 Hz). As empirical bands in each group shared the same quantity of bands, but displayed shifted frequency boundaries, we next examined individual variation and group differences in spatiotemporal properties.

**Figure 3:**
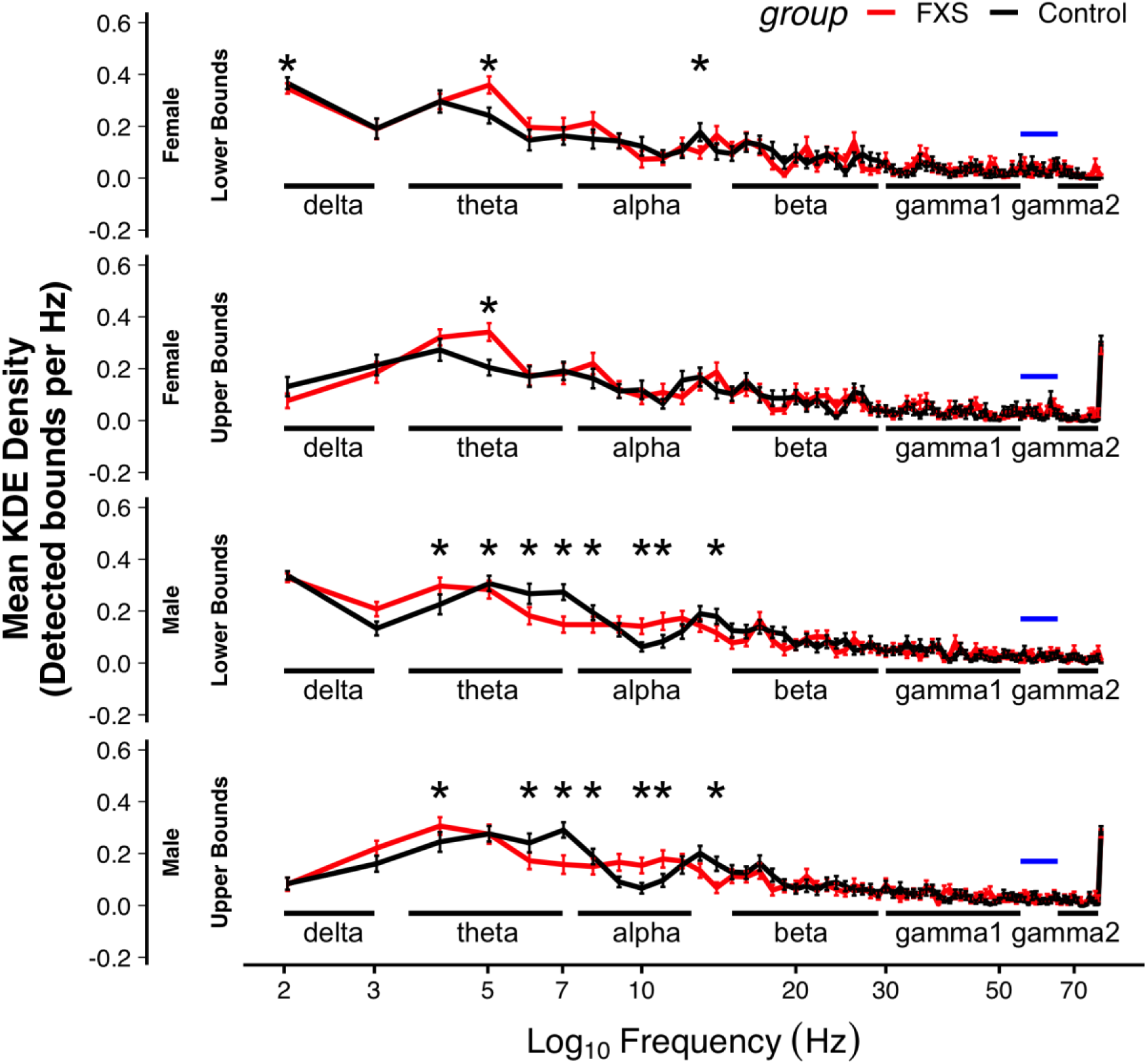
Group comparison. C. Mean kernel density estimates (KDE) of empirically defined frequency boundaries, separated by group and sex. Higher KDE values represent greater density of detected boundaries per frequency step. Lower and upper bounds were modeled and are visualized separately. A three-way interaction effect between Group X Sex X Frequency was present (Lower: F77, 10087=1.72, p=.005; Upper: F77, 10087=1.3, p=.047). Asterisks indicate statistical difference between control and FXS groups. Black bars provide contextual comparison with canonical frequency bands. Blue bars indicate line noise notch filter.

#### Spatiotemporal uniformity within detected boundaries

The squared correlation coefficient maps (**see Figure 2**) which define each detected empirical band represent the uniformity of spatiotemporal activity within a cluster. Overall, no strong correlation between the strength of the spatiotemporal correlation coefficient and the central frequency of each cluster was found (**see Supplemental Figure 1**). A weak inverse correlation was found in control females such that higher frequency bands had reduced correlation relative to lower frequency bands (r=-.15, p <.01).

#### Spatiotemporal changes in detected empirical alpha band

We defined each individual’s empirical alpha band as the detected cluster with the lower frequency bound closest too, but not exceeding 10 Hz (Cohen, 2021). **Figure 4** shows the lower and upper boundaries of individual empirical alpha band by group. The mean detected alpha band ranged in FXS from 7.6 to 13.0 Hz and in the control group from 7.8 to 12.9 Hz. Values in this range correspond approximately to the canonical alpha band. However, in contrast to canonical frequency bands, the empirical alpha band detected by GED involves a temporal (frequency boundaries) and spatial (topography) component.

**Figure 4:**
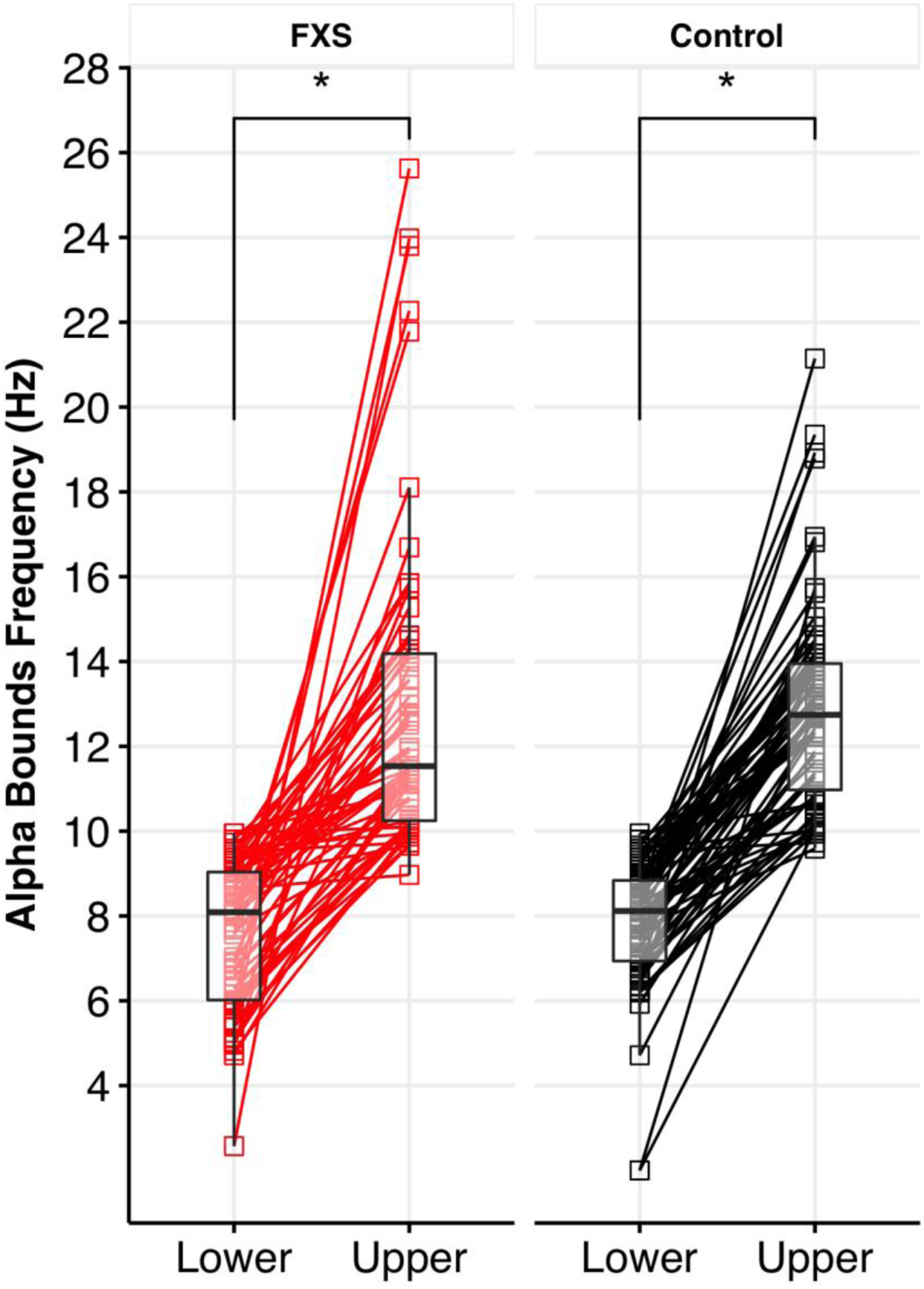
Analysis of empirical alpha component. A. Subject level paired plot of identified lower and upper alpha band boundaries for FXS and control groups. Each alpha band was identified as the closest, but not exceeding, lower bound to 10 Hz. Asterisks indicate significant within-group difference between lower and upper bound.

#### Frequency boundaries of empirical alpha band

We used an LME to examine if detected alpha boundaries varied based on group, sex, or boundary type (upper or lower). The results found a significant main effect of boundary type (F_1,131_=275; p<.001), but no effect of group or sex (**see Supplemental Table 3**).

#### Extracting empirical alpha band topography

PCA-averaged eigenvector (*w*) of the detected alpha band represents a spatial filter (n-channels in length) that maximally separates alpha range S_MATRIX_ and the broadband R_MATRIX_. When *w* is applied to the alpha range S_MATRIX_ (*w*^T^S_MATRIX_), we can plot the resulting component vector (with length equal to the number of channels) as a scalp topography. Though some of these components may be non-physiological (i.e., noise), certain frequencies will display topographical projections which resemble cortical dipoles of source projections (Haufe et al., 2014). We found at the subject level, the empirical alpha band consistently resulted in anatomically interpretable topography (**see Supplemental Figure 2**).

#### Assessing distribution of alpha band topography

As each empirical alpha band represents a distinct spatiotemporal configuration, different dipolar projections may represent different anatomical origins(Delorme et al., 2012; Zuure et al., 2020). Specifically, the reduced canonical alpha power and increased theta and gamma power reported in FXS resembles the EEG motif of thalamocortical dysrhythmia seen in certain other neuropsychiatric conditions(De Ridder et al., 2015; Llinas et al., 1999; Vanneste et al., 2018). A defining feature of TCD syndromes is a “slowed” alpha rhythm which has an anterior topography. We predicted that in FXS, the empirical alpha band would also display an anterior topography. We calculated spatial variance (r^2^) to correlate the topography of the empirical alpha band for each subject with either an ideal anterior or posterior template (**see Figure 5**)(Zuure et al., 2020). Variance, rather than correlation, is used in this scenario as the eigenvectors produced by GED have uncertainty in sign.

**Figure 5:**
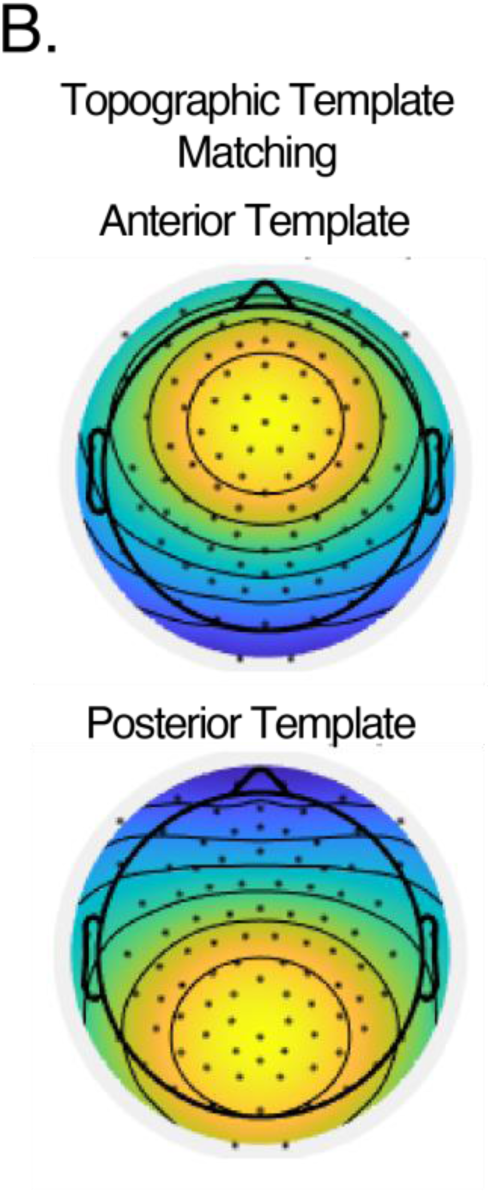
B. Matching templates. EEG templates were created with gaussian map centered on electrode FCz or PCz. Templates were used to determine similarity of neural projection to either anterior or posterior topography.

We conducted an LME to examine effects of group, sex, and template type (anterior or posterior) on r^2^. A two-way interaction effect of group and template type was present (F_1,131_=4.9, p=.028; **see Supplemental Tables 4-6**). First, between group post-hoc contrasts found that the empirical alpha band in FXS was more closely matched to the anterior template compared to controls (Anterior Template: FXS r^2^-Control r^2^, M=.10, 95%CI [.02,.18], F=2.36, p=.02, **see Figure 6**). Furthermore, within group contrasts found a significant preference for posterior template of the empirical alpha component in controls (Anterior r^2^-Posterior r^2^: M=-.18, 95%CI [-.27, -.10], F=-4.19, p<.001), but not FXS participants (Anterior r^2^-Posterior r^2^: M=-.05, 95%CI [-.14, -.04], F=-1.12, p=.26; **see Figure 7**.

**Figure 6:**
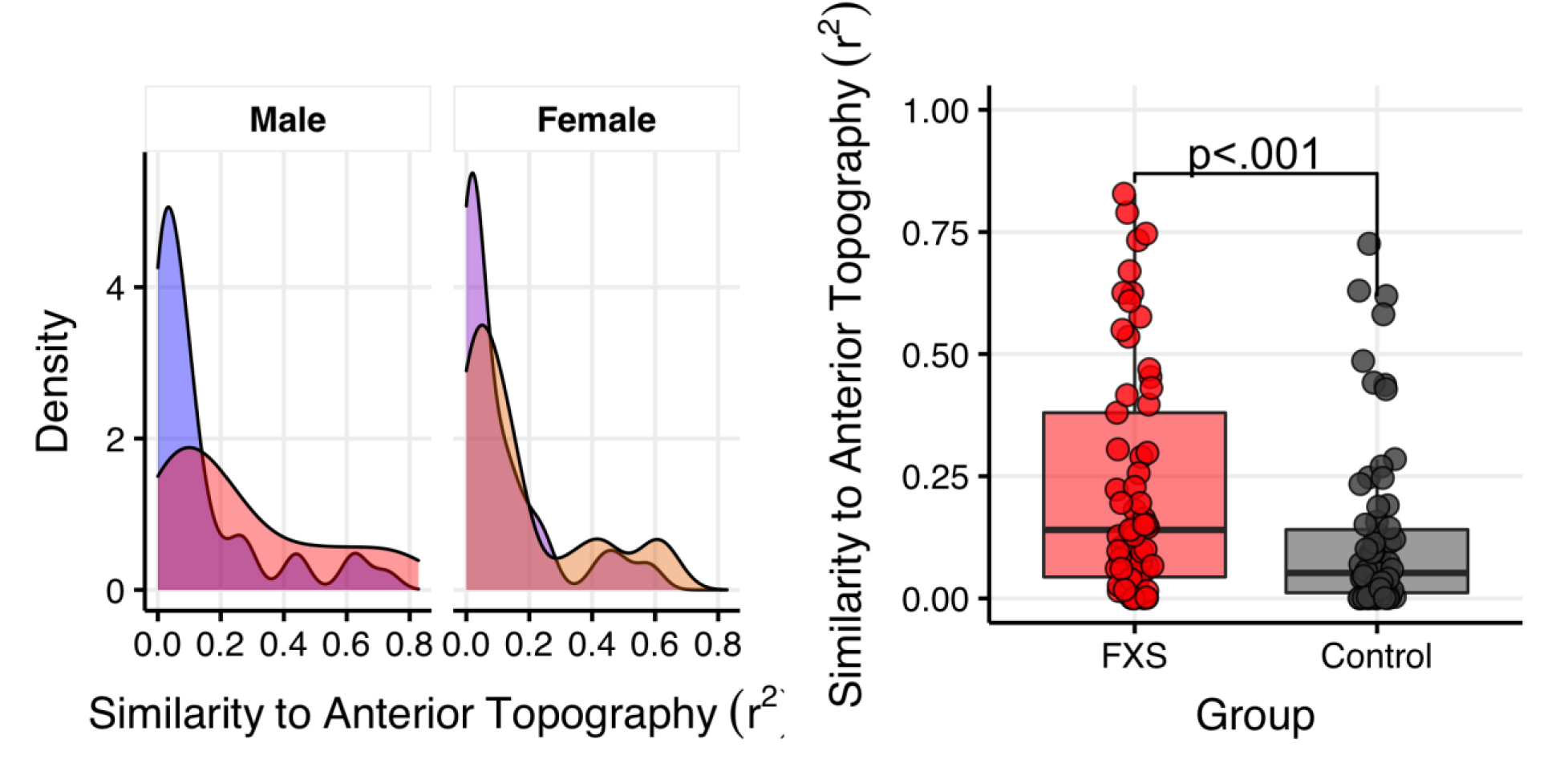
Left. Anterior topography preference for alpha component in FXS. Density plot of correlations representing subject-level matching of identified alpha component and mid-frontal topology. Right. Topography of alpha component is significantly altered in FXS participants (F1,131=4.9, p=.028). Alpha component demonstrates a greater degree of similarity to a mid-frontal distribution.

**Figure 7:**
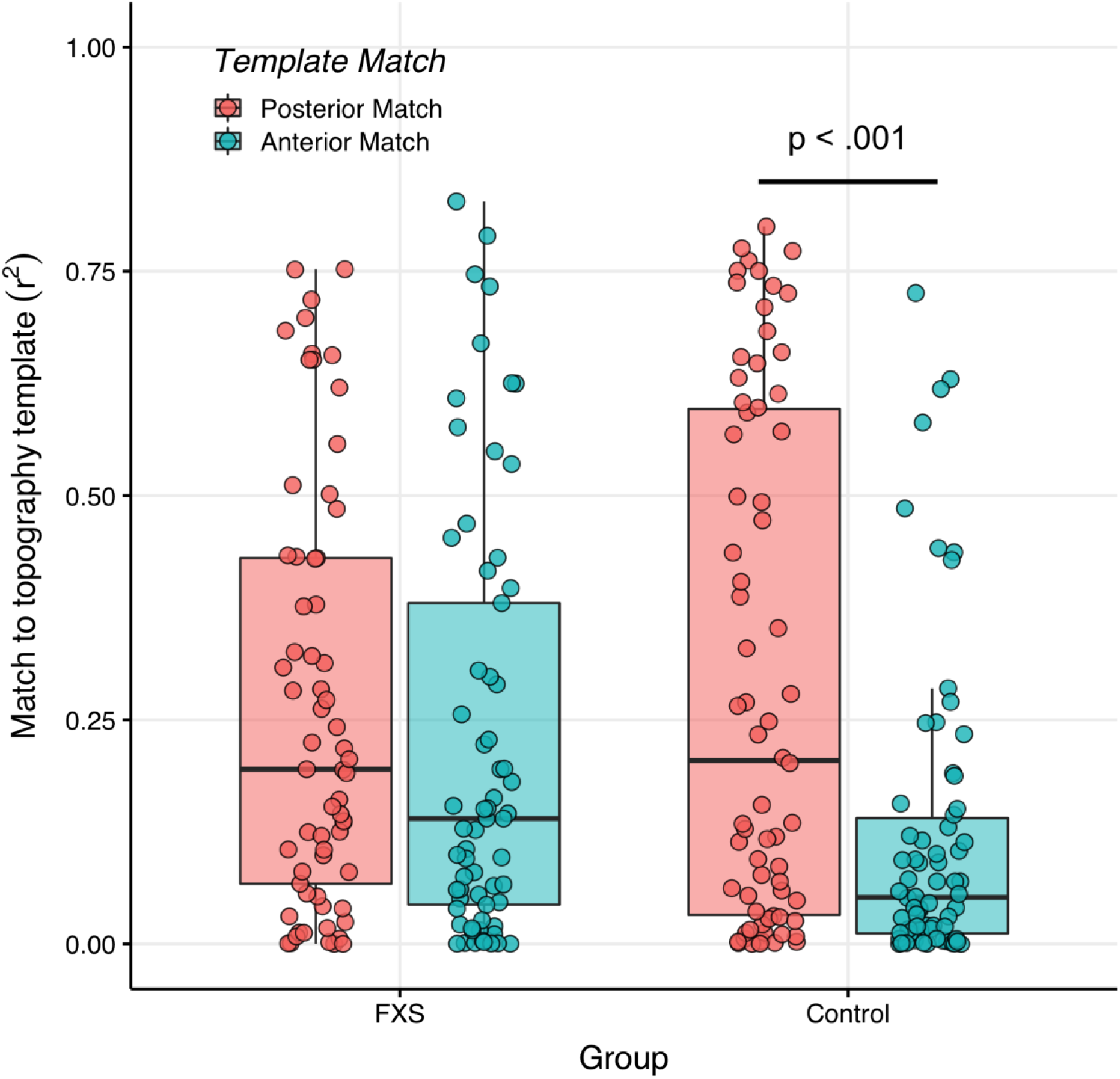
Posterior topography preference for alpha component in controls. Boxplot depicting similarity of empirical alpha component to either anterior or posterior topography. Scatter plot (with random horizontal offset for clarity) shows subject-level data. Comparing topography within groups revealed a lack of differentiation among participants with FXS, but a posterior topography preference among controls participants.

#### Empirical Alpha Band Power is Correlated with Auditory Attention in FXS

Following identification of altered topography of alpha component participants affected by FXS, age-adjusted clinical correlations were explored. The male-only subset was considered *a priori* as they exemplified a subset with minimal to no FRMP expression. We performed Pearson’s correlations of intelligence subscales, measures of neuropsychiatric symptoms, and an auditory attention task commonly used in FXS research (see Methods for details) with derived features from their individualized alpha component. In males with FXS, following 5% FDR correction and controlling for age, auditory attention was positively correlated with absolute alpha power (r_partial_=.57, p<.001, n=32, 5% FDR p < .05, **see Figure 8** for simple correlation plot). No clinical correlations emerged associated with topography. Full table of uncorrected correlations is presented in **Supplemental Table 7 and 8**.

**Figure 8:**
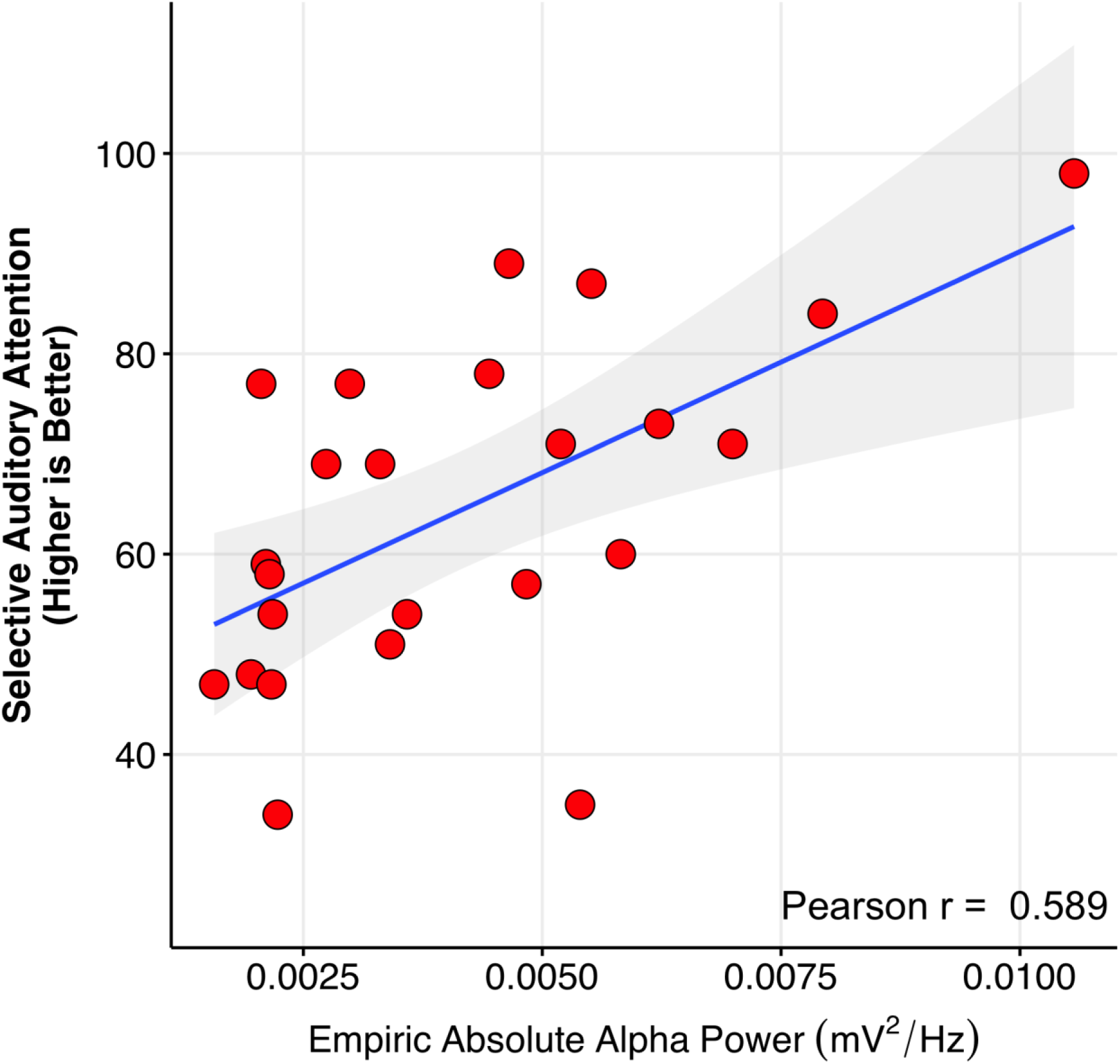
Clinical correlation: Higher performance in auditory attention was positive correlated with increased relative alpha power in males with FXS. Alpha power was empirically derived with subject level boundaries visualized in Figure 2A. Simple correlation plot presented for visualization only. Adjusted age-corrected results presented in text (r_partial_=.57, p<.001, n=32, 5% FDR p < .05).

## Discussion

Alpha oscillations are among the most prominent and physiologically relevant EEG patterns in the human brain. Marked alterations in alpha oscillations have been reported in FXS, including a leftward shift (lowering) of peak alpha frequency (PAF) and decreased global alpha power (Smith et al., 2021; Van der Molen & Van der Molen, 2013; J. Wang et al., 2017). It remains unclear, however, whether alpha activity differs between FXS and controls in terms of the frequency range it covers or its spatial configuration. Using a data-driven approach, we used spatiotemporal to classify patterns of neural activity from resting-state EEG into empirically-defined frequency bands. We found participants with FXS displayed: 1) similar total quantity of empirical activity bands as controls, 2) a “slower” dominant rhythm with a redistribution of lower-frequency boundaries, 3) an anterior topography of the empirical alpha band, and 4) significant clinical correlation of empirical alpha band activity with neurosensory dysfunction. This study clarifies key aspects of the nature of alpha oscillations in FXS and raises novel hypotheses related to the role of thalamocortical function in causing the heightened neural excitability in FXS, which is largely unknown.

### Incorporating spatial dimension to frequency band classifications

The quantity of detected empirical activity bands did not differ between FXS and control groups. Thus, both groups had similar total divisions of neural activity, though with key differences in the upper and lower frequency limits and the spatial topography of these empirically-defined bands. Compared to six to eight canonical frequency bands commonly reported, the quantity of detected empirical frequency bands for each group was approximately twelve. There is a likely reason for this increase: empirical activity bands model different spatial configurations within a frequency range. For example, multiple empirical bands with distinct topographies may fall within the range of a single canonical frequent band. This can be clearly seen in the subject-level tracings (**see Figure S2**). In both groups, fewer spatiotemporal coherent bands were identified at greater than 30 Hz, in which narrow-band activity may not sufficiently separate from background signal. In terms of alpha band activity, the mean lower (7.5 Hz) and upper (13 Hz) boundaries of the empirical alpha band in both FXS and control groups were remarkably consistent with the canonical alpha band. The present findings support the delineation of brain activity by frequency bands, as commonly performed with canonical frequency bands. However, the addition of a spatial dimension to account for variability in EEG activity may enhance the ability to model heterogeneity and provide insight physiology.

Various forms of data-driven analysis of EEG data have been successfully employed to model heterogeneity in neurodevelopmental populations. For example, across participants with neurodevelopmental conditions, a decrease in individual peak alpha frequency (IAPF) has been reported (Dickinson et al., 2018; Dickinson et al., 2019; Wang et al., 2013; Jun Wang et al., 2017).

### Altered distribution of empiric alpha frequency boundaries in FXS

Participants with FXS have a higher density of empirical band frequency boundaries in the canonical theta range, and in males with FXS, a sharp reduction of boundaries in the canonical alpha range. This suggests, especially in males with FXS, a leftward shift or “slowing” of the dominant frequency from alpha to theta frequency range (**see Figure 1C**). Narrow-band activity of spectral peaks is thought to reflect the compound synchrony of thousands to millions of neuronal currents. Based on the comparable number of empirical bands between the groups, these results point to a redistribution of neural activity towards lower frequencies.

### Altered configuration of empiric alpha topography in FXS

Spatially, alpha oscillations in FXS had a more anterior projection than in controls. In addition, within the FXS group, the distribution of alpha oscillations in FXS lacked the posterior-to-anterior gradient seen in healthy individuals. This analysis defined empirical alpha bands as the range of coherent spatiotemporal activity around 10 Hz. This approach ensured we captured a frequency typically defined within the canonical alpha band, with the addition of individualized lower and upper boundaries. The isolated empirical alpha components we analyzed represent a source projection, rather than scalp alpha activity. GED when applied to a narrow band and a broadband signal matrix is a form of linear source separation. The topographical plots, though reminiscent of spectral power plots, more accurately represent areas in which GED minimizes commonalities (e.g., volume conduction) and boosts differences between signals. The final topographies will capture noise or non-neural activity but can also effectively capture dipole components of brain activity. In this sense, these findings may indicate alterations in alpha source generators, which can be observed as spatiotemporal changes in the empirical alpha band.

### Association of empirical alpha activity with sensorineural function

In individuals with FXS, particularly in males, the auditory attention task of the Woodcock-Johnson III was associated with absolute “individualized” alpha power. The auditory attention task does not rely on caregiver reports and measures speech-sound discrimination and resistance to auditory stimuli(McGrew & Woodcock, 2006). We speculate that alpha power may serve to index behaviors in which thalamocortical pathways may take a central role, such as in perceptual or attentional tasks(Bollimunta et al., 2011; Foxe & Snyder, 2011). No other behavioral correlations survived multiple comparisons and may indicate that higher-order cognition and behavioral reports may not be easily modeled by a single EEG measure.

### Evidence for thalamocortical dysrhythmia in FXS

The slowing and shift of alpha activity from posterior to anterior electrodes have been previously reported with modulation of thalamocortical networks via anesthetic drugs and in pathological states (Vijayan et al., 2013). These features along with increases in gamma power associated with cross-frequency coupling (CFC), have been described as a neurophysiological motif called thalamocortical dysrhythmia (TCD). TCD was first described in magnetoencephalography tracings from neuropsychiatric conditions including depression, epilepsy, neurogenic pain, tinnitus, and Parkinson’s disease(Llinas et al., 1999). Thus far, electrophysiological findings drawn from FXS are consistent with TCD dynamics including elevated gamma activity (Ethridge et al., 2016; Van der Molen & Van der Molen, 2013; J. Wang et al., 2017). In essence, evidence from invasive recordings and theoretical models suggest that TCD disorders have alterations in cortical and thalamic synchrony leading to alterations of thalamocortical drive and regulation of alpha oscillations(De Ridder et al., 2015; Martire et al., 2019; Vanneste et al., 2018).

### Consequences of alpha-band alterations

What significance would spatiotemporal changes in low-frequency activity have on neural function in FXS? Theta and alpha activity transverse the cortex and have been associated with cognitive, emotional, and physiological functions(Zhang et al., 2018). These “traveling waves” are thought to coordinate activity, including binding cell assemblies, multiplex information, and selectively process data(Jensen & Mazaheri, 2010; Klimesch, 1999; Nunez et al., 2001). For example, in the visual system, the alpha phase can modulate the efficiency of visual information processing and alpha amplitude is associated with the activity of task-relevant and task-irrelevant regions(Busch & VanRullen, 2010). Furthermore, activity within the theta-alpha spectrum is commonly associated with cross-frequency coupling of higher frequency rhythms (i.e., gamma activity) and includes heavy contributions from subcortical sources, especially the thalamus(Canolty & Knight, 2010; Lega et al., 2016; Roux et al., 2013). Thus, one reasonable consequence of major alterations in low frequencies may be reduced efficacy of system level operations which display a frequency preference(Buschman et al., 2012; Canolty & Knight, 2010).

### Limitations

Use of GED necessitates specifying parameters that can change the results(Cohen, 2021). Despite a systematic approach, varying the search step size (epsilon) can alter band detection. In addition, other methodological decisions such as filtering approaches and artifact cleaning may also change the result. To minimize potential selective bias, our preprocessing was blind to the diagnosis group, and we applied the analysis parameters uniformly across groups. Readers can also view each data-driven tracing and access the entire dataset and computer code to reproduce the current results and explore alternative parameters.

### Conclusions

Genetically, individuals with FXS share a common cause, but vary greatly in phenotypic manifestations (including sex and mosaic effects). This difference may be due to contribution varying gene expression, compensatory processes, or stochastic factors (Baker et al., 2019). FMRP-deficient circuits have significantly altered electrophysiological properties(Goswami et al., 2019; Jonak et al., 2020; Lovelace et al., 2018; Lovelace et al., 2020). Here we used a “bottom-up” approach to empirically model spatiotemporal patterns of brain activity in a large sample of participants with FXS(Cohen, 2021). EEG is being used to investigate the links between physiology and behavior and measure target engagement of interventions in NDCs (Ewen et al., 2019; Sahin et al., 2018). Here we provide compelling evidence that even within a monogenetic clinical population, coherent patterns of spatiotemporal activity reside within EEG data and contribute to additional variability to group-level analysis. In addition, empirical methods may have increased sensitivity to within-subject designs, such as clinical trials to assess individual responses. Such findings may broaden therapeutic opportunities, including novel neural targeted “pacemaker” approaches to optimize endogenous rhythms. Such limits impede treatment discovery as drugs leading to recovery of biological and behavioral functions in Fmr1-/-KO mice have not shown efficacy in clinical trains in humans(Erickson et al., 2018; Harkins et al., 2017).

## Methods

### Participants

Resting-state EEG data were collected from participants with FXS (n=65; 37 males, 28 females) and age- and sex-matched typically developing controls (n=70; 41 males, 29 females). Six other invididuals recruited were not included in analyses: two for insufficient valid data epochs (2 FXS), three for excessive line-noise artifact (1 FXS and 2 controls), and one due to intolerance of EEG acquisition (1 FXS).

### Participant Consents

Study procedures were approved by the Cincinnati Children’s Hospital Medical Center Institutional Review Board. Participants provided written informed consent (or assent as appropriate) before participation.

### Clinical Diagnosis and Measures

Southern Blot and polymerase chain reaction confirmed diagnosis of FXS. Participants with FXS were excluded if they had a history of unstable seizures (any treated seizure within one year) and scheduled use of benzodiazepines. Controls were free of history or treatment neuropsychiatric illness as reported via clinical interview. Intellectual function was assessed using the Stanford-Binet Intelligence Scale 5th Ed. (SBS)(G, 2003). To capture cognitive variability and avoid floor effects, deviation intelligent quotient (IQ) scores were computed (Sansone et al., 2014). Primary caregivers completed assessments for FXS patients, including the Social Communication Questionnaire (SCQ)(Rutter et al., 2003), Anxiety, Depression, and Mood Scale (ADAMS)(Esbensen et al., 2003) and Woodcock-Johnson III Tests of Cognitive Abilities Auditory Attention subscale(McGrew & Woodcock, 2006).

### Dataset Availability

Data is available to the public as federally mandated at the National Database for Autism Research (NDAR).

### EEG Acquistion and Processing

Five-minutes of spontaneous resting-state EEG data was acquired at 1000 Hz sampling rate with a 128-channel geodesic electrode net (HydroCel) using an EGI NetAmp 400 amplifier (Magstim, Eugene, OR). As in previous studies, participants were seated comfortably while watching a silent video (standardized across participants) in order to facilitate cooperation with procedures (J. Wang et al., 2017). Preprocessing (**see Supplemental Methods**), including filtering and artifact correction, was performed using EEGLAB (14.1.2) and custom MATLAB (2018a) scripts (http://github.com/cincibrainlab/vhtp). EEG preprocessing was blinded to group assignment, participant identification, and study date. Eye, cardiac, and myogenic artifacts were removed via independent component analysis on each dataset using an extended INFOMAX algorithm (Lee et al., 1999; Mullen et al., 2015). For each subject, one-hundred and twenty seconds of artifact free data (sixty, 2-second epochs) were analyzed.

#### Source separation by Generalized Eigenvalue decomposition

To generate empirical frequency components we implemented generalized eigenvalue decomposition (GED) (Cohen, 2021) and a clustering algorithm to create spatiotemporal components from continuous EEG data (**Supplemental Methods**). First, we constructed channel covariance matrices to capture the linear relationships between pairs of channels and their corresponding amplitude time series. GED was performed between sequential finite-impulse bandpass filtered signal (S) matrices between 2 to 80 Hz in 500 logarithmically spaced steps and the broadband signal (R) matrix for each subject.

GED, in this context, is a method of source separation that maximizes the separation between two channel covariance matrices while ignoring common features. With GED, spatial patterns of underlying sources can be derived by minimizing volume conduction and spatial autocorrelation in contrast to univariate electrode analysis (e.g., spectral power topography). An empirical frequency range can then be determined by clustering spatial-temporal components produced by GED. The aim of this procedure is to identify frequency bands (e.g., alpha band activity) in an individual participant basis using spatial topography and oscillation frequency simultaneously. This approach can be useful for individualizing frequency bands for each participant rather than assuming canonical bands, and so that case control differences in frequency bands can be identified and related to clinical features.

#### Subject-level Clustering of Frequency Components

For each subject, the results of sequential GED between S and R matrices generated a frequency correlation matrix that represent coherent spatiotemporal activity over consecutive frequencies (**Figure 1A)**. To generate individual frequency band characterization, a non-hierarchical clustering method, density-based spatial clustering (dbscan), was implemented to search and define each cluster. The number of clusters was not predefined. Instead, the value of epsilon, which is the granularity of the resolution in searching for neighborhoods, was optimized for each subject based on a systematic quality function (Cohen, 2021). For each cluster, we identified the precise lower and upper empirical frequency boundary. Next, for each subject, we smoothed the lower and upper boundary vectors applying a kernel density estimator (KDE; 2 Hz full-width half-maximum Gaussian kernel). With this approach, a continuous likelihood estimate of true frequency boundaries was created, allowing for averaging of data across subjects and comparisons between groups.

#### Derivation of alpha source components

With a primary focus on alpha band activity, each participant’s empirical alpha component was defined as the spatial-temporal cluster whose lower boundary is closest, but does not exceed 10 Hz, and the corresponding upper boundary. Spectral power: Absolute power (μV^2^/Hz) was computed for the derived alpha component time series using Welch’s power spectral density (using MATLAB *pwelch*. Relative power was defined as the proportion of absolute alpha power in the total absolute spectral power of the broadband signal. Topography: To capture the degree of similarity of the spatial topography if empirical alpha components, we calculated shared spatial correlation between two “ideal” 128-channel topographic templates by applying a Gaussian centered at electrode FCz (“Anterior preference”) and PCz (“Posterior preference”) (**see Figure 2B**) (Zuure et al., 2020). As the alpha component has uncertainity in sign, the resulting correlations were squared to obtain shared spatial variance (r^2^).

### Statistical Testing

Statistical analyses were performed in R (version 4.1). Significance was set at p < .05 for all tests and adjusted for multiple comparisions using a 5% false discovery rate(Benjamini & Hochberg, 1995). Group comparisions of participant demographics, QC metrics, and component number were compared using one-way analysis of variance ANOVA.

To examine qualitative differences in frequency component boundaries and alpha component topography, multi-level linear mixed effects models (LME) were conducted using the *lme4* package (Bates et al., 2014). In each model, the subject was included as a random effect. Model details are presented adjacent to each result. Post-hoc contrasts of least-square means, with 5% false discovery rate corrected p-values for significance testing, were calculated using the *emmeans* package. Partial correlations were used to examine age-corrected relationships between clinical and neurophysiology features with 5% FDR p-value adjustment.

## Data Availability

All data produced in the present study are available upon reasonable request to the authors

## Acknowledgments

We thank the participants and families who participated in this study. The present study was federally funded by the National Institutes of Health (NIH) Fragile X Centers (U54HD082008 and U54HD104461).

